# Predicting COVID-19 spread and public health needs to contain the pandemic in West-Africa

**DOI:** 10.1101/2020.05.23.20111294

**Authors:** Beaugard Hémaho Taboe, Kolawolé Valère Salako, Calistus N. Ngonghala, Romain Glèlè Kakaï

## Abstract

The novel coronavirus (COVID-19) pandemic is causing devastating demographic, social, and economic damage globally. Understanding current patterns of the pandemic spread and forecasting its long-term trajectory is essential in guiding policies aimed at curtailing the pandemic. This is particularly important in regions with weak economies and fragile health care systems such as West-Africa. We formulate and use a deterministic compartmental model to (i) assess the current patterns of COVID-19 spread in West-Africa, (ii) evaluate the impact of currently implemented control measures, and (iii) predict the future course of the pandemic with and without currently implemented and additional control measures in West-Africa. An analytical expression for the threshold level of control measures (involving a reduction in the effective contact rate) required to eliminate the pandemic is computed. Considering currently applied health control measures, numerical simulations of the model using baseline parameter values estimated from West-African COVID-19 data project a 60% reduction in the daily number of cases when the epidemic attains its peak. More reduction in the number of cases will be achieved if additional public health control measures that result in a reduction in the effective contact rate are implemented. We found out that disease elimination is difficult when more asymptomatic individuals contribute in transmission or are not identified and isolated in a timely manner. However, maintaining a baseline level of asymptomatic isolation and a low transmission rate will lead to a significant reduction in the number of daily cases when the pandemic peaks. For example, at the baseline level of asymptomatic isolation, at least a 53% reduction in the transmission rate is required for disease elimination, while disease elimination is also possible if asymptomatic individuals are identified and isolated within 2 days (after the incubation period). Combining two or more measures is better for disease control, e.g., if asymptomatic humans are contact traced or identified and isolated in less than 3 days then only about a 31% reduction in the disease transmission rate is required for disease elimination. Furthermore, we showed that the currently implemented measures caused the time-dependent effective reproduction number to reduce by approximately 37% from February 28, to August 24, 2020. We conclude that COVID-19 elimination requires more control measures than what is currently being applied in West-Africa and that mass testing and contact tracing in order to identify and isolate asymptomatic individuals early is very important in curtailing the burden of the pandemic.

## 1. Introduction

A new strain of coronavirus (SARS-CoV-2), that emerged from Wuhan, China is the cause of the COVID-19 pandemic that is currently ravaging the world [1–3]. As of May 12, 2020, about 4, 337,602 confirmed COVID-19 infections and 292,451 deaths were reported worldwide [4–7]. Although the pandemic originated from China, most of the reported cases are now from the United States of America (approximately 1,408,636 infections and 83,425 deaths). The epicenter of the pandemic is expected to shift to sub-Saharan Africa, which as of May 12, 2020 had reported about 46,976 confirmed cases and 1,111 deaths. Of these, about 19, 750 confirmed cases and 428 deaths were from West-Africa. The first confirmed West-African case was in Nigeria on February 27, 2020, i.e., approximately two months after the first case was officially announced in China [1]. The highest burden of the disease in West-Africa on May 12, 2020 is in Ghana(with about 4, 700 cases and 22 deaths) and Nigeria (with about 4,641 cases and 150 deaths) [8]. A multilayered-risk assessment based on nine risk factors identified Nigeria as the West-African country with the highest COVID-19 risk [9]. Martinez-Alvarez et al. [10] projected that some West-African countries, e.g., Burkina Faso and Senegal might experience sharp increases in the number of COVID-19 cases that are similar to those observed in European countries in March and April, 2020.

Humans can acquire the novel coronavirus when they come into contact with contaminated surfaces or from droplets released by infectious symptomatic and asymptomatic individuals [11]. Mild to moderate infection symptoms of the disease include fever, cough, sore throat, nasal congestion, malaise, headache, muscle pain, and shortness of breath (or tachypnea in children). In severe cases, fever is associated with severe dyspnea, respiratory distress, and tachypnea [12]. Currently, there is no vaccine or widely accepted drug for COVID-19. Therefore, governments and individuals are forced to rely on public health preventive measures such as basic hygiene, travel or movement restrictions, social-distancing, wearing masks, etc. Current control measures being implemented in West-Africa include regular hand washing with hydroalkolic solutions, quarantine of suspected cases, isolation of confirmed cases, social distancing (e.g., travel restrictions, school closures, and banning gatherings involving more than 10 people), contact tracing, and testing and treating identified cases. Additionally, wearing of masks in public was recently recommended as another control measure in many countries in this region. Unfortunately, it is difficult to implement these basic public health measures effectively in some West-African countries due to wide-spread poverty and poor investment in health care (staff, equipment, and infrastructure). For example, in many West-African countries less than five hospital beds and less than two medical doctors are allocated to every group of 10,000 inhabitants [13]. Furthermore, the annual per capita spending on health for about half of the countries in the region is less than US$50, i.e., about 50 times below the average per capita health spending in countries like Italy and Spain-two European countries with high COVID-19 burden [14]. As a result of poor investment in health care and health care systems, the region has been an epidemic hotspot for emerging and neglected tropical diseases including the Ebola virus (EBOV), Lassa virus (LASV), and Buruli ulcers in recent years [15, 16]. With such inadequate healthcare systems and personnel in the region, the impact of COVID-19 might be catastrophic. To compound matters, some countries in West-Africa are still not well prepared to tackle this pandemic despite the observed devastating impacts and trend of the pandemic in the US and Europe [10].

There is, therefore, the need to exploit every existing tool and/or develop new tools that can be useful in guiding public health decision-making in the fight against the pandemic. This includes developing and using new qualitative and quantitative methods such as mathematical models. There has been an influx of mathematical models to assess the impact of COVID-19 and to guide public health response in different cities, countries and regions of the world (see, for example, [17–27]). Few of these mathematical model frameworks have focused on the COVID-19 pandemic situation in West-Africa as a whole. Among these few models, some have focused only on one or two countries, e.g. Nigeria and Ghana, while others have used used basic regression or exponential growth models [25, 26] to analyze and predict disease trends. Martinez-Alvarez et al. [10] used a simple graph-plot based comparative analysis of observed COVID-19 data for the first 23 days of the pandemic to examine the future trajectory of the epidemic in six West-African countries (Burkina Faso, Senegal, Nigeria, Cote d’Ivoire, Ghana, and The Gambia). This far, only one study has considered the entire region of West-Africa [27]. In this study, a co-variate-based instrumental variable regression framework was used to predict the number of disease cases (2.8 million by June 30, 2020) and to assess the epidemiological, socio-economic and health system readiness of the region to the pandemic.

In this study, we develop a new susceptible-exposed-infectious-recovered (SEIR)-type mathematical model for COVID-19 in West-Africa and use it to (i) study the current transmission pattern of COVID-19, (ii) evaluate the impact of currently implemented public health control measures such contact tracing, social distancing and use of face masks on COVID-19 transmission, and (iii) predict the future course of the pandemic with and without currently implemented and additional control measures in West-Africa. In addition to incorporating various basic public health control measures in our model, it accounts for asymptomatic infectious individuals – a class of individuals that render disease control more difficult since they do not exhibit clinical disease symptoms, although they contribute to disease transmission [21]. To our knowledge, this is the first study that has used an explicit SEIR-type compartmental mathematical model together with data for the West-African pandemic period (from February 28, 2020 to April 23,2020) to understand the spread of the COVID-19 pandemic in the region, assess the impact of current control measures, and discuss possible further control measures that might be required to better control the pandemic in the region.

## 2. Methods

### 2.1. Model framework

We develop a deterministic *SEIR-type* model framework, where the entire population of West-Africa is subdivided in four categories: Susceptible (*S*), Exposed (*E*), Infectious asymptomatic (*I_a_*), infectious symptomatic (*I_s_*), infectious at treatment or isolation centres (*I_c_*), and Recovered (*R*). The model accounts for public health control measures that lead to a reduction in the force of infection λ, such as social distancing and face mask use. Following the approach in Ngonghala *et al*. [17], we model the force of infection as (1 − Ψ)(*β_a_I_a_* + *β_s_I_s_* + *β_c_I_c_*)/(*S* + *E* + *I_a_* + *I_s_* + *R*), where *β_k_* (*k* ∈{*a, s, c*}) is the disease transmission rate by individuals in the *I_k_* class and 0 ≤ Ψ ≤ 1 is the percentage reduction in disease transmission due to public health control measures such as social distancing and face masks use. Note that Ψ=0 implies that no public health control measure leading to a reduction in disease transmission is applied and that Ψ=1 implies that implemented public health control measures are efficient enough to stop disease transmission. After the average incubation period 1/*σ*, a proportion *π* of the exposed population does not develop symptoms, i.e., joins the *I_a_* compartment, while the remaining proportion (1 − *π*) join the *I_s_* class. Identification and isolation of infectious asymptomatic individuals, e.g., through contact tracing occurs at rate *ρ_a_*, i.e., 1/*ρ_a_* is the average time it takes for an infectious asymptomatic individual to be identified and isolated, while infectious asymptomatic individuals develop symptoms at rate *α*. Isolation or hospitalization of symptomatic cases occurs at rate *ρ_s_*, i.e., 1/*ρ_s_* is the average time it takes for an infectious symptomatic individual to be isolated. Individuals from the *I_a_,I_s_*, and *I_c_* compartments recover at rates *γ_a_*,*γ_s_*, and *γ_c_*, respectively, while individuals in the *I_s_* and *I_c_* classes die due to COVID-19 at respective rates *δ_s_* and *δ_c_*. The flow diagram for the model system is presented in Fig. 1.

**Fig. 1:**
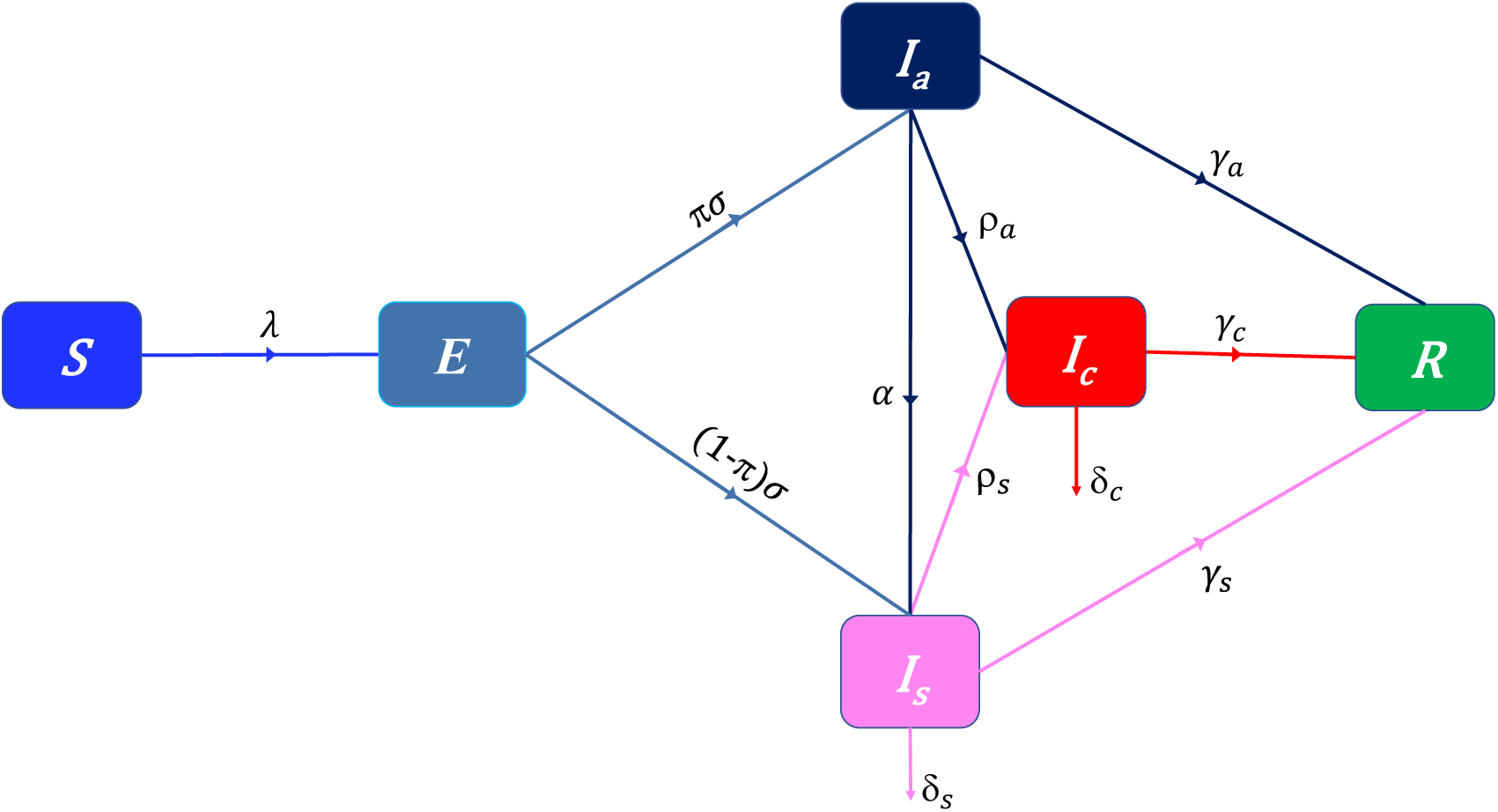
Flow-chart of COVID-19 model showing the flow of humans between different compartments. The susceptible population is denoted by *S*, the exposed population is denoted by *E*, the infectious asymptomatic population is denoted by *I_a_*, the infectious symptomatic population is denoted by *I_s_*, the isolated infectious population is denoted by *I_c_*, and the recovered population is denoted by *R*. The parameters of the model are described in the text.

Using the flow-chart and the preceding description, we obtain the following model system:

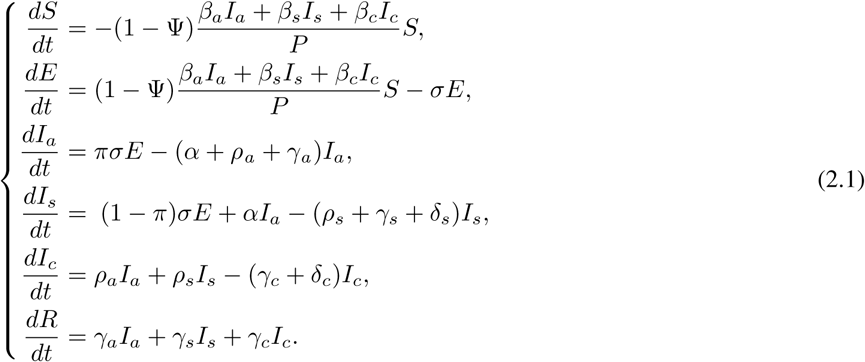

### 2.2. West-African COVID-19 data and parameter estimation

West-Africa comprises sixteen countries: Benin, Burkina-Faso, Cape Verde, Ghana, Guinea, Ivory Coast, Mali, Mauritania, Niger, Nigeria, Senegal, Togo, Sierra Leone, Liberia, Guinea-Bissau and Gambia. Based on the latest United Nations estimates, the current population of West-Africa is 399, 386, 502, i.e., about 5.16% of the world population [28]. The region includes nine of the 25 poorest countries in the world [29]. Application of basic public health control measures against COVID-19 spread in most West-African countries started on March 15, 2020, i.e., about 17 days after the outbreak in the region. Since mass testing in the region only started around April 24, 2020 and since many countries in the region are not carrying out mass testing yet, we avoid using data collected after April 23 in our study. Therefore, the data considered for this analysis spans the period from February 28, 2020, i.e., one day after the first case through to April 23, 2020 (before mass testing started), i.e., 39 days after the start of control measures. This allowed us to assess the dynamics of the disease before the start of control measures (i.e., the worst case scenario) and for the entire period (with control measures) by estimating model parameters for the first 17 days of the outbreak, when no control measures were put in place and then for the entire period of data collection (Feb. 28-Apr 23). In West-Africa, the reported confirmed cases are infectious symptomatic individuals who have been tested at a treatment center or infectious asymptomatic individuals who have been contact-traced and then tested at a treatment centre. Hence, we assume that confirmed disease cases correspond to individuals in the *I_c_* class, i.e., isolated infectious individuals at isolation or treatment centers. We use the cumulative number of cases in West-Africa, downloaded from the Global Rise of Education website [8] to fit our model and estimate some of the key parameters presented in Table 1. The fitting was done using a nonlinear least squares procedure, and involved finding the best set of parameter values that minimized the sum of the square differences between the observed number of daily cases in West-Africa and the predicted cumulative number of cases from our model. We repeated this procedure 5,000 times to approximate the distribution of each of the parameters by a normal distribution and obtain the mean value with a 95% confidence interval. The goodness of fit of our model, assessed using the Root Mean Squared Error (RMSE) was 2.25 and 70.62, respectively, for the fits before the onset of interventions and for the entire period of data collection. The estimated parameters are presented in Table 1 and the model fit is illustrated in Fig. 2 (a), (b). The other model parameters that were extracted from the literature or estimated based on COVID-19 information from West-Africa are presented in table 2. It is worth noting that the choice of parameters to estimate from the data or to extract from the literature was motivated by its importance to the transmission dynamics of the disease. Although the transmission rate at the treatment centre *β_c_*, is important in influencing disease dynamics, we extracted its value from the literature based on the experience in other infectious disease management (e.g: Ebola and Lassa fever) in West Africa. We considered the value in [30]. Fig. 2 (c), (d) illustrates the evolution trend of the predicted cumulative number of asymptomatic (*I*a), symptomatic (*I_s_*), and isolated infectious (*I_c_*) using the baseline parameters. This figure suggests that irrespective of the public health measures put in place, under-reporting of the number of disease cases is very high.

**Table 1:** Estimated model parameter values for the two periods (initial: February 28 -March 15, 2020 and entire: February 28 -April 23, 2020)

| Parameter | Value (Initial period) | CI (Initial period) | Value (Entire period) | CI (Initial period) |
| --- | --- | --- | --- | --- |
| $\beta_a$ | 0.7594 | [0.7312-0.7877] | 0.6080 | [0.5680-0.6480] |
| $\beta_s$ | 0.9724 | [0.9626-0.9821] | 0.6300 | [0.5929-0.6672] |
| $\rho_a$ | 0.0471 | [0.0457-0.0484] | 0.0369 | [0.0273-0.0464] |
| $\rho_s$ | 0.0040 | [0.0015-0.0065] | 0.0526 | [0.0393-0.0659] |
| $\gamma_s$ | 0.0116 | [0.0101-0.0131] | 0.0950 | [0.0879-0.1022] |
| $\delta_s$ | 0.0809 | [0.0712-0.0905] | 0.3079 | [0.2894-0.3264] |
| $\Psi$ (baseline) | - | - | 0.2518 | [0.2302-0.2734] |

**Fig. 2:**
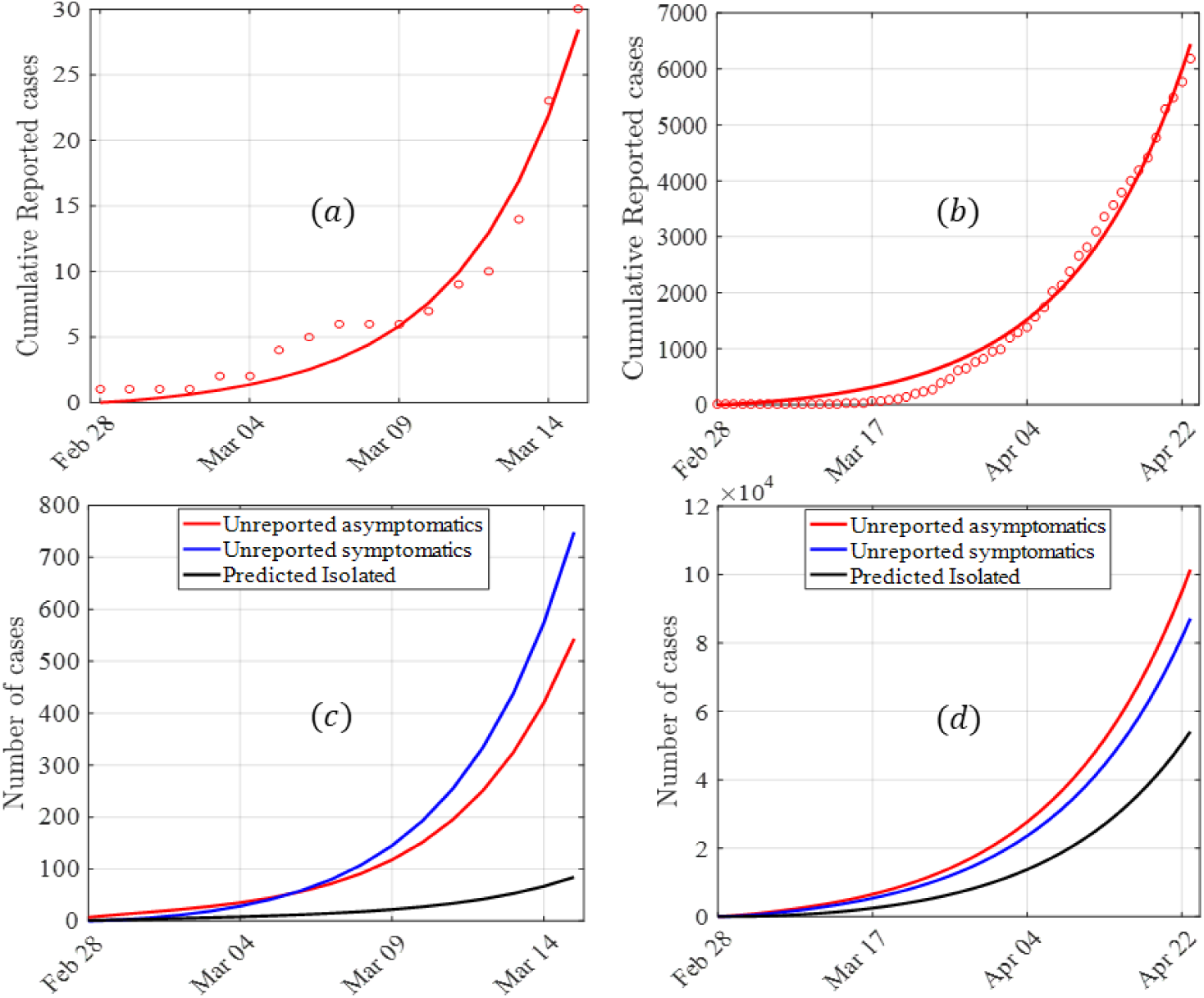
Model fit and evolution trend of reported and unreported disease cases. (a) Model fit for the period from February 28-March 15, 2020, i.e., the first seventeen days when no control measures were applied. (b) Model fit for the period from February 28-April 24, 2020. (c) Evolution of predicted number of asymptotic, symptomatic and isolated infectious cases period from February 28-March 15, 2020. (d) Evolution trend of predicted number of asymptotic, symptomatic and isolated infectious cases for the period from February 28-April 24, 2020. The initial conditions are *S* = 399, 386, 502; *E* = 1042; *I_a_* = 14; *I_s_* = 2; *I_c_* = 1; *R* =0 and the parameters used for the simulations are presented in Tables 1 (entire period) and Table 2.

## 3. Results

### 3.1. Analytical results

In this section, we derive the reproduction number of the model (which is defined as the average number of new infections generated by a typical infectious individual introduced in a population where everybody is susceptible) denoted by *R_c_* and a threshold value for the reduction in disease transmission required for eliminating the disease. Using the next generation matrix approach in [35, 36], the reproduction number is:

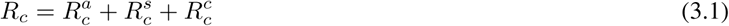

where

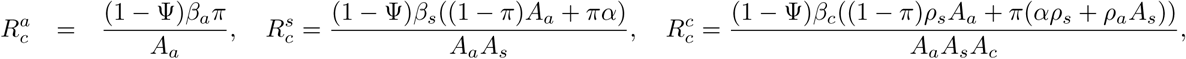

*A_s_* = *ρ_s_* + *γ_s_* + *δ_s_*,*A_a_* = *α + ρ_a_ + γ_a_* and *A_c_ = γ_c_ + δ_c_*. Observe from Eq. 3.1 that the reproduction number *R_c_*, is the sum of three components, which represent the contribution of the infectious asymptomatic class 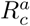, the infectious symptomatic class 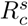, and the infectious isolated class, 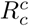. The time-dependent effective reproduction number is *R_c_S*/(*S + E + I_a_ + I_s_ + R*).

The approach in [35, 36] also assures us that the disease-free equilibrium 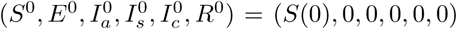 of system (2.1) is locally and asymptotically stable whenever the reproduction number *R_c_* is less than one and unstable if *R_c_* > 1. To derive the threshold reduction in the disease transmission Ψ*_c_*, required to reduce the control reproduction number to one, we set *R_c_* =1 in Eq. 3.1 and solve for Ψ. This yields:

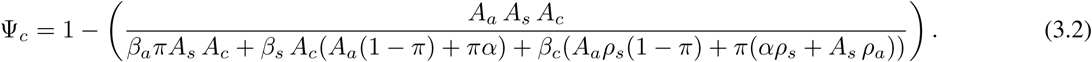

Note that the fraction in the right hand side of the threshold value Ψ*_c_* of Ψ is greater than zero and less than or equal to one since 0 ≤ Ψ ≤ 1. Note also that Ψ=Ψ*_c_*, which corresponds to the case in which *R_c_* =1 now serves as a threshold for an outbreak or the disease dying out. In particular, disease elimination is possible if Ψ > Ψ*_c_* (since *R_c_* < 1) and impossible if Ψ < Ψ*_c_* (since *R_c_* > 1). The epidemiological implication of this is that massive adherence to control measures that reduce COVID-19 transmission, e.g., social distancing can lead to disease elimination, while low adherence to such measures will reduce disease burden, but might not lead to elimination.

### 3.2. Numerical simulation results

#### 3.2.1. Current patterns of COVID-19 transmission

In this section, we use the parameters presented in Tables 1 and 2 to simulate system (2.1) and to investigate the impact of control measures on key model parameters and outputs such as the control and effective reproduction numbers *R_c_* and *R_e_*, respectively. The basic reproduction number *R*_0_, and the control reproduction number *R_c_* computed using this baseline parameter regime in Tables 1 and 2 are 2.09 (*CI* = [1.77 − 2.8]) and 1.60(*CI* = [1.45 − 1.75]), respectively. The control reproduction number corresponds to an approximately 23% reduction in the basic reproduction number due to implemented control measures. This calls for more effective control measures to limit the spread of the disease.

**Table 2:** Fixed parameters values for the two periods.

| Parameter | Value | Reference |
| --- | --- | --- |
| $\beta_c$ | 0.02 | [30] |
| $\delta_c$ | 0.0654 | Calculated from our data |
| $\gamma_a$ | 1/9.5 | [31] |
| $\gamma_c$ | 0.0206 | Calculated from our data |
| $\sigma$ | 1/5.1 | [32] |
| $\alpha$ | 0.304 | [33] |
| $\pi$ | 0.78 | [34] |

Fig. 2(c), (d) shows a growing number of unreported asymptomatic and symptomatic infectious individuals. Therefore, irrespective of the implemented public health control measures, there are still exponentially growing numbers of unreported or non-isolated asymptomatic and symptomatic infectious individuals. More simulations were carried out using the estimated parameters for the period from February 28-April 23, 2020 (Table 1) and the other parameters in Table 2 to test the short-term predictive power of the model (Fig. 3). Observe that the predicted cumulative number of cases resulting from the simulations captures the actual cumulative reported cases for the period from April 23 to April 30, 2020 very well.

**Fig. 3:**
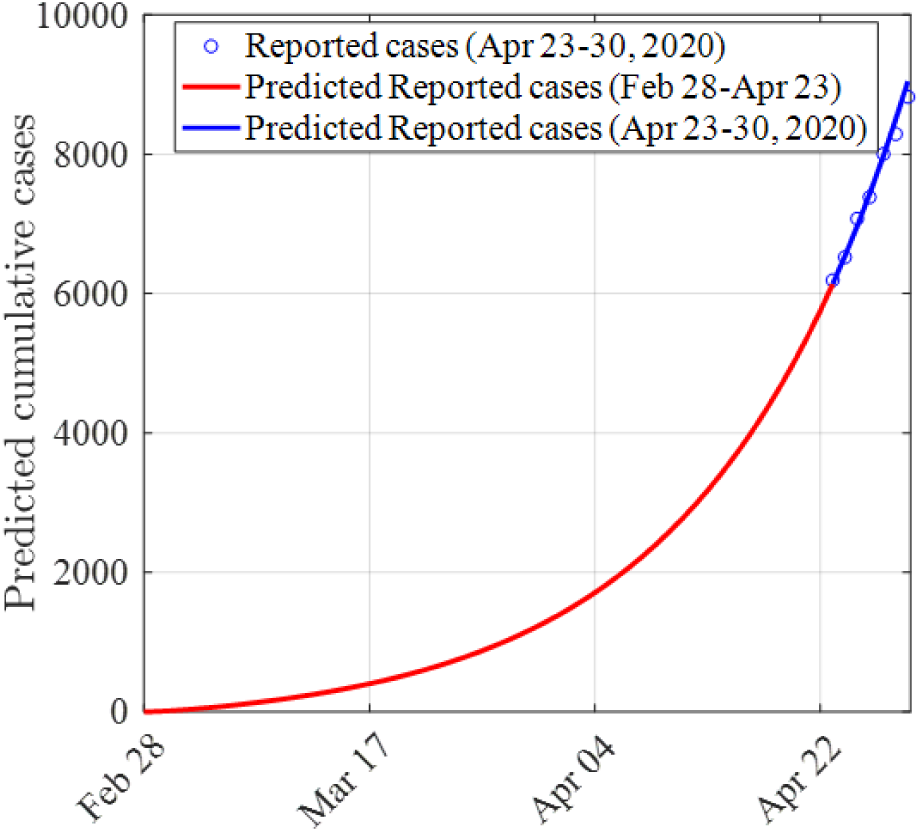
Short term Prediction of the cumulative number of COVID-19 cases in West-Africa. The initial conditions are *S* = 399, 386, 502; *E* = 1042; *I_a_* = 14; *I_s_* = 2; *I_c_* = 1; *R* =0 and the parameters used for the simulations are presented in Tables 1 (entire period) and Table 2.

### 3.3. Potential impacts of different health control measures on COVID-19 dynamics in West-Africa

Fig. 4 (a) shows the change in the control reproduction number in response to changes in the percentage reduction (Ψ) of disease transmission due to public health control measures such as social distancing or using face masks. It shows that about a 53% reduction in the disease transmission is required to reduce *R_c_* to one (Fig. 4 (a)). This value (53%) corresponds to the numerical value of Ψ*_c_* from Eq. (3.2) computed using the baseline parameter values from tables 1 and 2 and a 28% increase from the 25% baseline value of Ψ. Our results also show that there will be a 52% reduction in the effective reproduction number from February 28, 2020 to May 06, 2020 if no control measures are applied, and a 15% reduction if there is a 45% reduction in disease transmission from February 28, 2020 to June 02, 2021, i.e., an additional reduction of 20% in Ψ (Fig. 4 (b)). Additional simulations were carried out to investigate the dynamics of the system (2.1) for different values of Ψ. The results show that applying control measures at the baseline level of 25% will bring about a 60% reduction in the number of cases (from the worst case scenario) when the pandemic peaks and also delay the peak to mid September 2020. Improved control measures resulting in 35% and 45% reduction in disease transmission will lead to reductions of 76% and 92%, respectively, in the number of confirmed cases when the epidemic attains its peak (Fig. 4 (c)). Thus, improved control measures that are related to reducing disease transmission have a significant effect in reducing the number of cases and flattening the epidemic curve–an outcome that will prevent the already weak and fragile health care systems in West-Africa from being overwhelmed by a high COVID-19 burden.

**Fig. 4:**
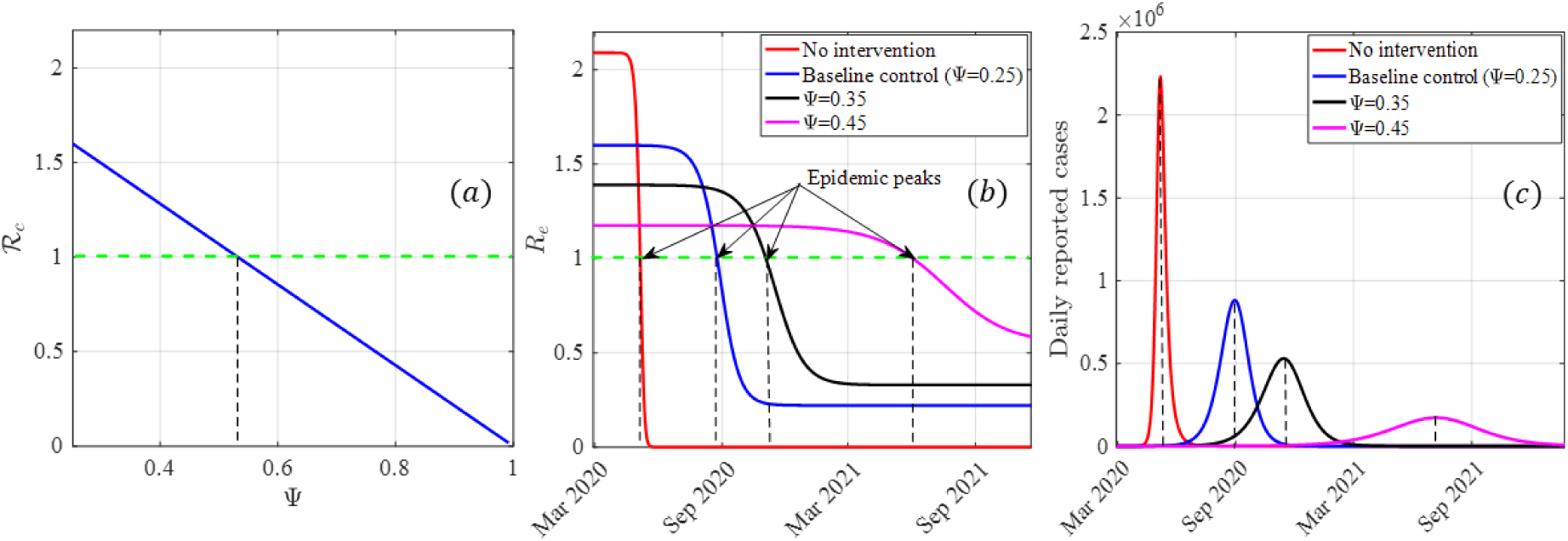
Impact of control measures assessed through a reduction in the disease transmission term on the control reproduction numbers and the epidemic peak. (a) Plot of the control reproduction number *R_c_* against the reduction in disease transmission due to the implementation of Ψ showing the threshold level of Ψ required to eliminate the disease. (b) Evolution of the effective reproduction number of COVID-19 in West-Africa for different values of Ψ. (c) Prediction and sensitivity of COVID-19 peak date to different values of Ψ. The initial conditions are *S* = 399, 386, 502; *E* = 1042; *I_a_* = 14; *I_s_* = 2; *I_c_* = 1; *R* =0 and the parameters are presented in Tables 1 and 2. The parameters for the initial period were used to plot the worst case scenario curves in the red (no control measure), while the parameters for the entire period were used for the other curves.

Other basic public health control measures include isolating symptomatic infectious humans and contact tracing and isolating asymptomatic humans. In our model we assume that isolating symptomatic infectious humans is at rate *ρ_s_* and contact tracing and isolating asymptomatic infectious humans is at rate *ρ_a_*. As of April 23, 2020, there was no mass testing in West-Africa. In particular, mostly suspected and symptomatic cases were tested. Hence, the most common pathway through which infectious asymptomatic individuals were identified in West-Africa was through contact tracing. Heat maps were plotted to investigate the individual and combined effects of pairs of control measures such as contact tracing, isolation, and using controls that lead to a reduction in disease transmission, e.g., mask use, social and physical distancing on COVID-19 in West-Africa (Fig. 5).

**Fig. 5:**
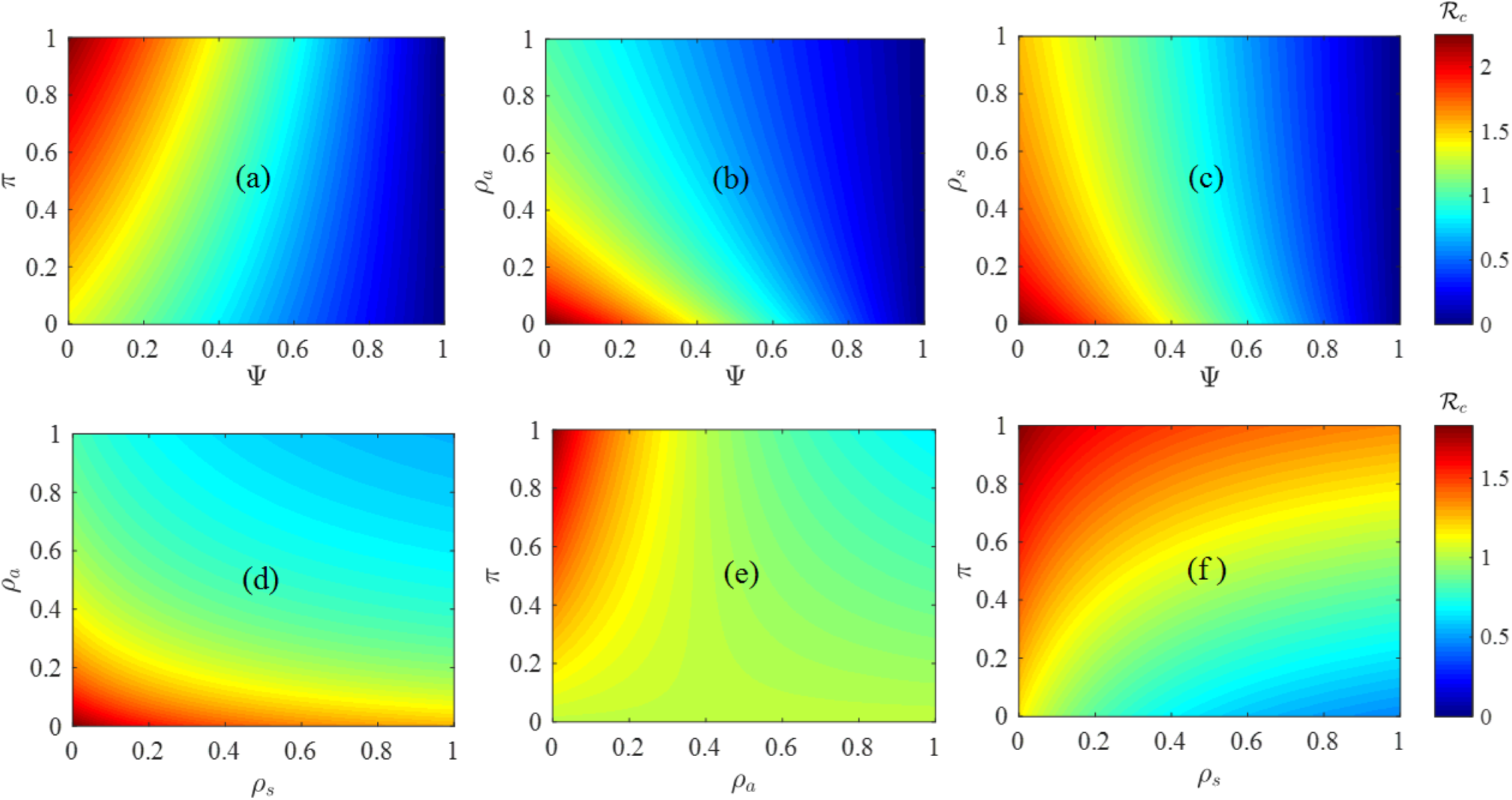
Sensitivity of COVID-19 control reproduction number *R_c_*, to different pairs of basic public health control measures.(a) Sensitivity of *R_c_* to the proportion of exposed humans who develop COVID-19 symptoms at the end of the incubation period (*π*) and the reduction Ψ, in the disease transmission rate.(b) Sensitivity of *R_c_* to the identification and isolation rate of asymptomatic infectious humans (*ρ_a_*) and Ψ.(c) Sensitivity of *R_c_* to the identification and isolation rate of symptomatic infectious humans (*ρ_s_*) and Ψ.(d) Sensitivity of *R_c_* to *ρ_a_* and *ρ_s_*. (e) Sensitivity of *R_c_* to *ρ_a_* and *π*. (f) Sensitivity of *R_c_* to *ρ_s_* and *π*.

Our analysis show that the spread of the disease decreases with a higher reduction in the disease transmission rate and a decreasing proportion of exposed humans who do not develop clinical disease symptoms (i.e., become asymptomatic) after the incubation period, *π* (Fig. 5 (a)). Thus, if a higher proportion of exposed humans do not develop clinical disease symptoms after the incubation period, a higher reduction in the disease transmission rate will be required to eliminate the disease. In particular, if every exposed individual develop symptoms at the end of the incubation period, i.e., *π* =0, then only a 29% reduction in disease transmission rate is required to eliminate the disease. But, if 78% and 50% of the exposed become asymptomatic after the incubation period, then 53% and 47% reductions in the disease transmission rate are required to eliminate the disease (Fig. 5 (a)). The combined impact of detection and isolation of asymptomatic humans (by contact tracing or mass testing) together with a reduction in the disease transmission rate is explored in Fig. 5 (b). If asymptomatic individuals are detected and isolated fast enough, e.g., within 2 days, i.e., *ρ_a_* =0.56, then a 20% reduction in the disease transmission rate would suffice for elimination, while if it takes a long time to detect and isolate asymptomatic individuals, e.g., within 10 days, a 48% reduction on the disease transmission rate is required to contain the pandemic in West-Africa (Fig. 5 (b)). We also found out that disease elimination is possible even if detection and isolation of asymptomatic humans is not complemented with any other measure. But such identification and isolation must occur in a timely manner, e.g., within 1 day, i.e., *ρ_a_* =1 (Fig. 5 (b)). On the other hand, disease elimination is also possible if more exposed humans develop symptoms at the end of the incubation period and asymptomatic humans are detected and isolated in a timely manner. For example, if only 78% of humans fail to develop symptoms at the end of the incubation period, then asymptomatic humans must be identified and isolated within 3 days for disease elimination to be possible (Fig. 5 (e)). Thus, disease control is more difficult if it takes long to contact trace and isolate asymptomatic humans.

Our results also show that timely isolation of symptomatic cases is important in reducing the disease burden in West-Africa, although disease elimination is only possible if isolation of infectious symptomatic cases is complemented with another control measure (Fig. 5 (c), (d)). In particular, if symptomatic humans are identified and isolated within 3 days, i.e., *ρ_s_* =0.39, then a 43% reduction in the disease transmission rate is required for disease elimination, while if it takes a long time to isolate symptomatic infectious individuals, e.g., within 10 days, a 51% reduction on the disease transmission rate is required to contain the pandemic in West-Africa (Fig. 5 (c)). When isolation of symptomatic infectious disease cases is complemented with identification and isolation of asymptomatic infectious cases, disease elimination is only possible through timely identification and isolation of asymptomatic cases (Fig. 5 (d)). Thus, identifying and isolating asymptomatic individuals in a timely manner contributes more in curtailing the pandemic in West-Africa than isolating symptomatic infectious individuals. Furthermore, if more exposed humans develop disease symptoms at the end of the incubation period and more symptomatic infectious individuals are identified and isolated in a timely manner, disease elimination is possible (Fig. 5 (f)). In particular, if 58% of exposed humans develop clinical disease symptoms at the end of the incubation period, then symptomatic infectious humans must be isolated within 2 days in order to eliminate the disease. However, if 37% of exposed humans develop clinical disease symptoms at the end of the incubation period, then symptomatic infectious cases must be isolated within 3 days in order to eliminate the disease.

## 4. Discussion

Mathematical models have been useful in understanding disease-outbreaks and in informing policy aimed at curtailing such diseases in a timely manner [17–19, 23, 24, 37]. In the context of the COVID-19 pandemic that is currently spreading around the world, mathematical models have been very useful in predicting the course of the disease and in assessing the impacts of basic public health control measures [17–19, 23]. In this study, we develop a mathematical model to inform the COVID-19 trend and possible course of control measures in West-Africa. The model is trained with COVID-19 data from West-African countries for the period from February 28, 2020 to April 23, 2020 and used to compute the reproduction number, as well as assess the impact of basic public health control measures in the region.

We obtained a reproduction number *R_c_* =1.60 for West-Africa, which is close to the reported value for South Korea (1.5) [38] but lower than the value reported for Wuhan one week before travel restrictions were introduced (2.35) [39]. Hence, although the disease is still spreading (*R_c_* > 1) in West-Africa, the spread is slow compared to other parts of world. This low transmission in the region can be attributed to a number of factors, e.g., the pandemic might still be in the early stage, since the statistics for West-Africa within the first seven weeks of the pandemic (3, 661 cases and 79 deaths), are similar to those of France and higher than those of the US (1, 025 cases and 28 deaths) when these countries were within the same outbreak time-frame. Also, the low transmission could be due to the fact that there is limited movement of individuals within the region and between the region and the rest of the world [9]. Furthermore, the low number of confirmed cases in West-Africa might result from under-reporting of cases as was the case in Peru [40]. This is supported by the growing number of asymptomatic infectious individuals observed in our study. De Leo et al. observed a similar trend in Mexico, where over 90% of the infectious population was asymptomatic [21]. In the absence of sufficient test kits in many West-African countries, and with the limitations in health care facilities and personnel, identifying cases is difficult [41]. Irrespective of the current trend, the COVID-19 pandemic is still a more serious health problem in West-Africa compared to previous outbreaks like the 2014 Ebola outbreak (1.50 ≤ *R_c_* ≤ 1.60) [42, 43] or the Lassa fever outbreak in Nigeria (1.29 ≤ *R_c_* ≤ 1.37) [44].

Although most countries in West-Africa started implementing basic control measures since March 17, 2020, our simulation results indicate that improvements are necessary to effectively control or eliminate the pandemic from the region. Because there is currently no safe and effective vaccine or drugs against the novel coronavirus, basic public health control measures have been used to curtail the pandemic in many parts of the world including West-Africa. These measures include contact tracing, isolation, and measures that lead to a reduction in disease transmission, e.g., mask use and social or physical distancing [17]. In the absence of such measures, the worst case scenario prediction for West-Africa from our model is over 2 million confirmed COVID-19 cases by mid May 2020 when the epidemic peaks. This projection is within the range (517, 489 15, 056, 314) in [27] for the same West-African region, although the peak in the latter study was predicted to occur in June 2020. The higher upper limit in [27] might be linked to the fact that the authors used linear models and considered the rate of infection between the first and second weeks of the epidemic in countries in which the number of cases might have been overestimated. We found out that basic public health control measures, especially those associated with a reduction in the disease transmission rate such as social distancing and mask use have a significant effect on reducing the burden of the disease, e.g, at baseline values, control measures reduce the number of cases when the epidemic peaks by about 60% and the disease transmission rate by about 25%. Also, improving control measures such about an estimated 45% reduction in the disease transmission is attained, reduces the number of confirmed cases when the epidemic attains its peak approximately 92%. Such interventions are useful in reducing ensuring that the already limited and fragile health care systems are not overwhelmed by the COVID-19 burden [19].

Our results suggest that early identification and isolation of both symptomatic and asymptomatic infectious individuals is an important step towards curtailing the burden of the pandemic. In particular, identifying and isolating asymptomatic individuals early enough, e.g., within 1-2 days after the end of the incubation period can result in disease elimination, even without any other control measure. The same is not true with identifying and isolating symptomatic individuals or when it takes a longer time to identify and isolate infectious asymptomatic individuals. In this case, the measure must be complemented with another measure that involves a reduction in the disease transmission rate in order to eliminate the disease. Asymptomatic infectious individuals can be identified through contact-tracing and testing. However, since mass testing in most West-African countries only started during the last week of April, the primary way in which asymptomatic individuals were identified in West-Africa during the study period was through manual contact tracing and then testing the contacts suspected to have the virus. One way in which contact tracing has been used successfully to identify and isolate both pre-symptomatic and asymptomatic infectious individuals is through mobile phone applications, see for example, Ferretti et al. [45]. We showed that a higher reduction in the disease transmission rate, e.g., through social distancing or using masks will be required to eliminate the disease if a higher proportion of exposed humans do not develop clinical disease symptoms after the incubation period. These results highlight the importance of early identification and of disease cases and are consistent with the results of previous studies on Ebola virus and Lassa Fever virus in West-Africa [43, 46]. The results are also consistent with those of a study by Hellewell et al. [47], who used simulated data to illustrate that contact tracing and isolation is highly effective in controlling the pandemic and can lead to disease elimination within 3 months. Although, contact tracing is already being implemented in West-Africa, the measure is not very effective due to the lack of resources and the high community transmission in the region. For example, as of April 8, 2020, about 4 in every 5 reported cases in the region resulted from community transmission [5]. Therefore increased compliance with measures like social distancing and mask use is very important for the region.

Since the disease can be transmitted from one human to the other through droplets from infected individuals that can travel though a few meters in air [23, 48], using masks and social distancing are very important in the fight against the pandemic. Masks play a dual role–they prevent acquisition of the virus by infected individuals and also prevent infectious individuals from spreading the virus [17]. Both using masks and social distancing are ongoing in many West-African countries. However, the effectiveness of these measures in reducing the spread of the virus is linked to the extent to which they are applied, the compliance level by public, and how long they will last [17, 21]. Our analyses show COVID-19 can be eliminated in West-Africa through social distancing and using masks if 45% of the population complies with the measures. This is consistent with recent results in [17, 23, 49], where strict compliance with social distancing and mask use can lead to disease elimination. Some West-African countries, e.g., Benin have implemented measures involving isolating some densely populated portions of the country, especially overcrowded urban areas with dilapidated sanitary facilities and limited basic amenities like clean water [28]. This has a positive effect on reducing the spread of the virus.

Finally, our study suggests that mass application of control measures coupled with appropriate testing and timely isolation of asymptomatic humans will go a long way in keeping disease numbers low. However, with the fragile economic conditions and health care systems in the region, interventions like complete lock-down as observed in most European countries for a couple of months can be more catastrophic than the disease itself. Therefore, the choice of interventions by public policy makers in the region should aim at balancing the prevention of the epidemic with the need for maintaining livelihoods and social cohesion.

## 5. Conclusion

Our study confirms the fact that the novel coronavirus (COVID-19) is highly contagious and that infectious humans who do not show clinical disease symptoms (asymptomatic or unreported cases) contribute significantly in disease transmission. This study also indicates that early identification of these unreported cases through contact tracing or mass testing, and then isolating individuals who test positive plays a significant role in diminishing the burden of COVID-19 in West-Africa. Furthermore, this study suggests that social distancing measures such as stay-at-home orders, closing educational institutions, limiting mass gatherings, etc., and using masks can reduce the spread of the disease significantly, with the possibility of disease-elimination if about one in every two people in West-Africa respect these control measures. To enhance, the study shows that implementing more than one measure at the same time is better for disease control, and that under current control measures, the disease might not be disappearing from the region any time soon, unless there are improvements in these control measures. We conclude that mass-testing, contact tracing and isolation of confirmed disease cases, as well as improvements to the other existing basic public health measures (e.g., social distancing and mask use) in the region, are required to better manage or eliminate the pandemic. Also, due to uncertainties and various disparities between the economies and health care systems of countries within the region, we conclude that country-level studies are necessary and will provide more insights into disease dynamics and control in the region.

## Data Availability

I declare that all data used in the manuscript are available online.

https://ourworldindata.org/coronavirus-source-data

## Acknowledgments

KVS acknowledges the support of the Wallonie-Bruxelles International Post-doctoral Fellowship for Excellence, Belgium (Fellowship # SUB/2019/443681). CNN acknowledges the support of the Simons Foundation, USA (Award #627346). RGK acknowledges the support from the African-German Network of Excellence in Science AGNES and the Humboldt Foundation, Germany. The Authors acknowledge assistance from Sacla Edmond, Kotanmy Brezesky, and Agbovoedo Robert for collecting the initial data used for the project.

